# Cognitive Trajectories in the Nine Months following Recent-Onset Major Depressive Disorder

**DOI:** 10.1101/2025.05.08.25327188

**Authors:** Alexandra Stainton, Caroline X Gao, Georgina D Thomas, Robert Hester, Shayden Bryce, Katharine Chisholm, Siân Lowri Griffiths, Lana Kambeitz-Ilankovic, Julian Wenzel, Carolina Bonivento, Paolo Brambilla, Mariam Iqbal, Theresa K. Lichtenstein, Marlene Rosen, Linda A. Antonucci, Eleonora Maggioni, Joseph Kambeitz, Stefan Borgwardt, Anita Riecher-Rössler, Christina Andreou, André Schmidt, Frauke Schultze-Lutter, Eva Meisenzahl, Stephan Ruhrmann, Raimo K. R. Salokangas, Christos Pantelis, Rebekka Lencer, Olga Bienek, Georg Romer, Udo Dannlowski, Alessandro Bertolino, Rachel Upthegrove, Dominic B Dwyer, Nikolaos Koutsouleris, Stephen J Wood, Kelly Allott, the PRONIA Consortium

**Affiliations:** Orygen, Melbourne, Victoria, Australia; Centre for Youth Mental Health, University of Melbourne, Melbourne, Victoria, Australia; School of Public Health and Preventive Medicine, Monash University, Melbourne, Australia; University of Melbourne, Melbourne School of Psychological Sciences, Melbourne, Australia; Alfred Mental and Addition Health, Melbourne, Victoria, Australia; School of Psychology, The University of Sussex, UK; Institute for Mental Health and Centre for Human Brain Health, University of Birmingham, Birmingham, UK; Department of Psychiatry and Psychotherapy, Faculty of Medicine and University Hospital of Cologne, Cologne, Germany; Faculty of Psychology and Educational Sciences, Ludwig-Maximilians University, Munich, Germany; Scientific Institute, IRCCS Eugenio Medea, Pasian di Prato, Udine, Italy; Department of Neurosciences and Mental Health, Fondazione IRCCS Ca’ Granda Ospedale Maggiore Policlinico, University of Milan, Milan, Italy; Department of Pathophysiology and Transplantation, University of Milan, Milan, Italy; Department of Psychology, Woodbourne Priory Hospital, Birmingham, UK; Department of Translational Biomedicine and Neuroscience (DiBraiN) - University of Bari “Aldo Moro” - Bari, Italy; Department of Electronics, Information and Bioengineering, Politecnico di Milano, Milan, Italy; Department of Psychiatry and Psychotherapy, University of Lübeck, Lübeck, Germany; Medical Faculty, University of Basel, Switzerland; Department of Psychiatry, Psychiatric University Hospital, University of Basel, Switzerland; Department of Psychiatry and Psychotherapy, Medical Faculty, Heinrich-Heine University, Düsseldorf, Germany; Department of Psychology, Faculty of Psychology, Airlangga University, Surabaya, Indonesia; University Hospital of Child and Adolescent Psychiatry and Psychotherapy, University of Bern, Bern, Switzerland; Department of Psychiatry, University of Turku, Turku, Finland; Department of Psychiatry, University of Melbourne, Carlton South, VIC, Australia; Florey Institute of Neuroscience and Mental Health, The University of Melbourne, Parkville, VIC, Australia; Monash Institute of Pharmaceutical Sciences (MIPS), Monash University, Parkville, Melbourne, Victoria, Australia; Institute for Translational Psychiatry, University of Muenster, Muenster, Germany; Department of Child Adolescent Psychiatry and Psychotherapy, University of Münster, Germany; Clinic for Child and Adolescent Psychiatry, Psychotherapy and Psychosomatic Medicine, University Hospital Muenster, Schmeddingstrasse 50, 48149, Muenster, Germany; Department of Psychiatry, University of Oxford, Oxford, United Kingdom; Birmingham Early Intervention Service, Birmingham Women’s and Children’s NHS Foundation Trust, Birmingham, UK; Department of Psychiatry and Psychotherapy, Ludwig-Maximilian-University, Munich, Germany; Institute of Psychiatry, Psychology and Neuroscience, King’s College London, UK; Max Planck Institute of Psychiatry, Munich, Germany; School of Psychology, University of Birmingham, Edgbaston, United Kingdom

## Abstract

**BACKGROUND:** Specific cognitive impairments are common in major depressive disorder, impacting functioning and quality of life. However, the longitudinal course of cognitive functioning in depression is unclear.

**AIMS:** This study aimed to determine the longitudinal course of cognitive functioning following a recent onset of depression, as compared to healthy controls.

**METHODS:** Healthy controls and individuals experiencing a recent onset of depression (within two years) were recruited across ten European sites, completing the baseline and nine-month follow-up clinical, demographic, and cognitive assessment for the “PRONIA” study.

**RESULTS:** The sample comprised 421 participants (depression, N=152; healthy controls, N=269) aged 15-40 years (M = 25.4, SD = 6.1; 55% female). Linear mixed effects models demonstrated baseline cognitive deficits in the depression group across most domains, except for visual memory, visuospatial working memory, and emotion recognition. Deficits in verbal learning and memory, attention, processing speed, mental flexibility, phonetic and semantic verbal fluency remained stable. Auditory verbal working memory showed a lag trajectory, with healthy controls improving at a greater rate than the depression group. Finally, sustained attention followed a catch-up trajectory, with baseline deficits normalising over time relative to healthy controls. In the depression group, most cognitive improvements were associated with reductions in depression symptomatology, except for verbal learning and memory, and verbal fluency. The catch-up trajectory of sustained attention was associated with reductions in depression.

**CONCLUSIONS:** Specific cognitive impairments can already be evident at the first episode of depression, but cognitive functions show differential longitudinal trajectories irrespective of depressive course. Tailored treatment addressing cognition should be provided early to promote cognitive health and functional recovery.

## Introduction

Cognitive functions including memory, attention, executive function, and processing speed can be impaired in Major Depressive Disorder (MDD) (1, 2). Such impairments can significantly impact functioning and quality of life (3), contributing to disease burden (4). Crucially, the timing, severity, and pattern of change are not well understood, particularly early in the illness. Previous literature has characterised cognitive impairment as state, trait, or scar, depending on whether impairments resolve, present prior, or worsen, relative to the course of depression (5). While the term “scar” implies permanent damage from a depressive episode, cognitive worsening may be the result of co-occurring depression risk factors rather than the depression itself. Such “scars” may also be dynamic and reversible (6). Work examining the long-term trajectories of cognition following MDD has been limited by key methodological constraints. First, a reliance on cross-sectional evidence from different samples at different stages of MDD (e.g., currently depressed, in a period of remission, or experiencing recurrent depression) (e.g., 1). Second, while some longitudinal studies have shown enduring cognitive deficits in MDD relative to Healthy Controls (HC) (1, 7), they primarily include participants with more enduring illness and few had follow-ups longer than 3-6 months (7). Finally, only a small number of studies have examined cognitive functioning early in the illness course, which has the advantage of avoiding potential clinical confounders such as chronic or recurrent course, long-term treatment, or substance use/abuse (5).

Five hypothetical cognitive trajectories have been proposed in the early course of mental illness (8), and can also be applied to the period following recent-onset MDD (ROD): 1) *No-deficit*, reflecting no impairment relative to HC at any time, 2) A stable *deficit* endures over time, 3) A *catch-up* trajectory reflects an impairment that diminishes over time, 4) A *lag* is characterised by a failure to improve at the same rate as HC, and 5) A *deterioration* is the most extreme negative trajectory, with performance worsening over time. These trajectories also need to be interpreted within the developmental context, because in younger individuals, cognitive improvement is expected even in the context of depression, but cognitive development varies by domain (9).

Very few studies have examined longitudinal cognitive performance following ROD (10–14).

The follow-up periods ranged from one month to five years and findings have been variable. Some identified enduring processing speed and executive function impairments (10–12). Other studies observed that baseline deficits in memory, attention, processing speed, working memory, mental flexibility, and inhibition improve over time, indicating a catch-up trajectory relative to HC (11, 13, 14). These studies have been limited by their own methodological constraints, including single measurement of HC performance, meaning there was no control for normative changes or practice effects (15). Some studies have limited analysis to ROD participants in remission at follow-up (11, 13, 14), thus assuming that all cognitive improvements are related to symptomatic remission. Finally, these studies have been limited by very small sample sizes at follow-up (10, 12), or by limited or complex cognitive measurements that are difficult to interpret and replicate.

The literature to date has not provided sufficient understanding of the trajectory of cognitive functioning in the early course of MDD. This study aimed to provide a definitive answer to this important question through rigorous methodology not utilised in previous studies. We aimed to conduct the largest study to date in this area, measuring a broad range of cognitive functions in individuals with ROD and HC at baseline and nine-month follow-up. Our previous study of this sample demonstrated baseline cognitive deficits in the ROD group relative to HC (16). For this longitudinal analysis, we hypothesised that for each cognitive function measured (a) performance of the overall study population would improve with time, and (b) performance of the ROD group would still be poorer on average than the HC group. Investigation of whether the rate of change over time differed between the two groups (i.e., group by time interaction) was central to the aim of this study. Owing to the limitations and mixed findings of previous work, a priori hypotheses about the trajectories of specific cognitive functions in the ROD group relative to HC were not specified. A secondary aim was to investigate whether change in cognitive performance was associated with reduction in depressive symptoms.

## Materials and Methods

### Design and Setting

Individuals with ROD (i.e., a first-episode of MDD, <2 years) and HC participated in the “PRONIA” (Personalised Prognostic Tools for Early Psychosis Management) study (17). The current analysis used demographic, clinical, and cognitive testing data that were collected at baseline and a nine (+/-three) month follow-up.

### Participants

Participants aged 15-40 years were recruited through ten international sites (Germany: Munich, Cologne, Münster, and Düsseldorf; UK: Birmingham; Italy: Udine, Bari, and Milan; Finland: Turku; Switzerland: Basel). Full PRONIA inclusion and exclusion criteria have been reported previously (17, 18), and criteria for the present study are presented in Supplementary Table 1 and Supplementary Figure 1.

### Measures

#### Clinical Assessment

Demographic and medication information were collected during a face-to-face clinical interview. The SCID-I/P was used to establish DSM-IV diagnosis of MDD (19). Self-reported depressive symptoms were measured using the Beck Depression Inventory version 2 (BDI-II), comprising 21-items (20). Total score was interpreted as minimal (0-13), or mild (14-19), moderate (20-28) or severe (29-63) depression severity.

#### Cognitive Assessment

Baseline intelligence quotient (IQ) was estimated from age-derived scaled scores of the Matrix Reasoning and Vocabulary subtests of the Wechsler Abbreviated Scale of Intelligence, second edition (WASI-II) (21). Longitudinal cognitive performance was assessed using a battery of twelve tests, each with one primary outcome score (see Supplementary Table 2 and previous reports (18, 22)).

### Procedure

Participants provided written informed consent (plus parent/guardian consent for those <18 years). All measures were completed at both timepoints. Tests were conducted in the participants’ native language and administered in a set order. Participants were financially reimbursed for study participation. PRONIA was approved by ethics committees at each recruitment location.

### Statistical Analysis

Data cleaning was conducted in Microsoft Excel and demographic variables were analysed in IBM SPSS Statistics, Version 30 (23). All other statistical analyses were conducted in R Studio (24).

#### Multi-level Modelling

Linear mixed-effects models with random intercepts (Multilevel Modelling, MLM) were used to assess whether the trajectory of performance on 12 cognitive tests (Table 1), differed between the ROD and HC groups (25). The trajectory differences were tested using a group-by-time interaction term. Models were fitted using the lmer function in the lme4 package (26) using Restricted Maximum Likelihood (REML) estimation, a recommended realistic and unbiased estimation technique (25). The model also controlled for sex and age as confounding factors.

**Table 1.** Baseline Demographics, Mean Duration of Follow-up and Subjective Assessments of Depression.

|  | Recent-Onset MDD group<br>(N = 152) |  |  |  |  | Healthy control group<br>(N = 269) |  |  |  |  | t | p-value |
| --- | --- | --- | --- | --- | --- | --- | --- | --- | --- | --- | --- | --- |
|  | N | M | SD | Min | Max | N | M | SD | Min | Max |  |  |
| **Age**, years | 152 | 25.5 | 6.1 | 15.1 | 40.9 | 269 | 25.4 | 6.0 | 15.0 | 40.0 | -0.277 | 0.782 |
| **IQ**, estimated | 144 | 105.6 | 11.8 | 68<sup>a</sup> | 128 | 252 | 108.7 | 9.9 | 78 | 133 | 2.610 | 0.010<sup>b</sup> |
| **Education**, years | 151 | 14.6 | 3.0 | 7.0 | 25.5 | 268 | 15.9 | 3.02 | 9.0<sup>c</sup> | 25.0 | 4.284 | <0.001<sup>b</sup> |
| **Test Interval**, weeks | 152 | 42.4 | 4.7 | 28.7 | 52.0 | 269 | 42.7 | 4.4 | 29.0 | 52.1 | 0.784 | 0.433 |
| **BDI-II** |  |  |  |  |  |  |  |  |  |  |  |  |
| T0 | 142 | 26.1 | 13.5 | 0 | 60.0 | 256 | 3.4 | 5.1 | 0 | 42.0 | -19.322 | <0.001<sup>b</sup> |
| T1 | 118 | 14.3 | 12.2 | 0 | 55.0 | 233 | 3.0 | 4.1 | 0 | 22.0 | -9.837 | <0.001<sup>b</sup> |
| Change (T1 - T0) | 111 | -11 | 12.4 | -51 | 15 | 229 | 0.2 | 4.1 | -15 | 18 | 9.268 | <0.001<sup>b</sup> |
| | N | n | % | | | N | n | % | | | $\chi^2$ | p-value |
| **Female** | 152 | 73 | 48 |  |  | 269 | 157 | 58 |  |  | 4.188 | 0.041<sup>b</sup> |

*Note.* BDI-II = Beck Depression Inventory edition 2; IQ = Intelligence quotient; MDD = Major depressive disorder. <sup>a</sup>One individual with an estimated IQ of 68 was retained for analysis as it was deemed that this still fell within the acceptable range given the standard error for measurement around true scores from the 2-subtest version. 95% confidence interval for full scale IQ estimate for the individual's age would fall within the range of 63-77(21). <sup>b</sup>Statistically significant at a level of $p \leq 0.05$ . <sup>c</sup>An outlier of 0 Education Years was removed from analysis.

Missing cognitive assessment and BDI-II data (N for each test presented in Table 2 and eTable3), were handled using multiple imputation via the Multiple Imputation by Chained Equations (MICE) (27). Data were imputed with a wide format, stratified by group (28), using the missForest package (29). The imputed datasets were then reshaped to a long format for analysis, with 25 imputations generated and results pooled using Rubin’s rules (30).

The diagnostic plots (e.g., Tukey-Anscombe plots and Q-Q plots) for MLMs suggested minor violations of the linearity assumption for models (31), mostly due to the skewed distribution of the random intercept. Hence, sensitivity analyses were conducted with the same models and were fitted with generalized additive model (GAM) using mgcv package, where random intercepts were modelled as a smoothed function (32). Further sensitivity analyses ran the original model scaled to the minimum and maximum scores of each cognitive test and also excluded individuals in the ROD group who had experienced multiple MDD episodes at follow-up.

The association between reductions in depressive symptom severity and improvements in cognitive test performance over the follow-up period was examined in the ROD group only.

Depressive symptom scores on the BDI-II were baseline-centred prior to multi-level modelling (as above, age and sex also included as confounders) to differentiate the within- and between-person associations (33). The model produced two coefficients: 1) the baseline BDI-II scores reflecting between-person variation; and 2) changes in BDI-II scores from baseline, representing the within-person effect.

## Results

### Study Sample

The final sample comprised 421 participants (ROD, N=152; HC, N=269) aged 15 to 40 years (M = 25.4, SD = 6.1; 55% female). Participant characteristics were broadly similar in the two study groups, though the HC group had significantly higher estimated IQ and education years and comprised a higher percentage of females (Table 1). The mean test interval was approximately 9.8 months in both groups. Average depression severity on the BDI-II was much higher in the ROD group than in HCs at both timepoints, with an average reduction from moderate to mild observed in the ROD group at follow-up. Additional clinical characteristics of the ROD group at baseline and follow-up are presented in Supplementary Table 3 of the online supplement.

### Cognitive Performance

Baseline cognitive performance of the broader PRONIA study have been published previously (16, 18, 22). For the present ROD and HC groups, cognitive test score means, standard deviations and ranges at baseline and follow-up are presented in Supplementary Table 4.

### Linear Mixed-effects Models

Data suitability for MLM was assessed prior to model fitting. Sample size at level two of the model (i.e., the number of participants) was above 300 for each cognitive test, considered suitable for MLM (34). The proportion of missing data for most cognitive tests was 5%, but notably higher for TMT-A (9%), TMT-B (10%) and RAVLT (17%). Based on Little’s Missing Completely at Random (MCAR) test (p<.001) missing data were not MCAR for any cognitive test. In general, the proportion of missing data was higher at follow-up compared to baseline, and in the ROD group compared to the HC group; except for RAVLT for which missingness was higher in HCs. Based on a Shapiro-Wilk test for each cognitive test, scores were not normally distributed (p<.001). Visually, histograms were broadly normal for cognitive tests, although skewness (positive and negative) and outliers were evident. Outliers in outcome variables were not excluded as there was no theoretical reason to exclude plausible test scores.

#### Cognitive Trajectories

Results from the MLM examining group, time, and group-by-time (cross-level) interactions are presented in Figure 1. To enhance interpretability, the predicted margins are plotted in Figure 2, illustrating the longitudinal trajectories of cognitive performance outcomes separately for the ROD and HC groups.

**Figure 1.**
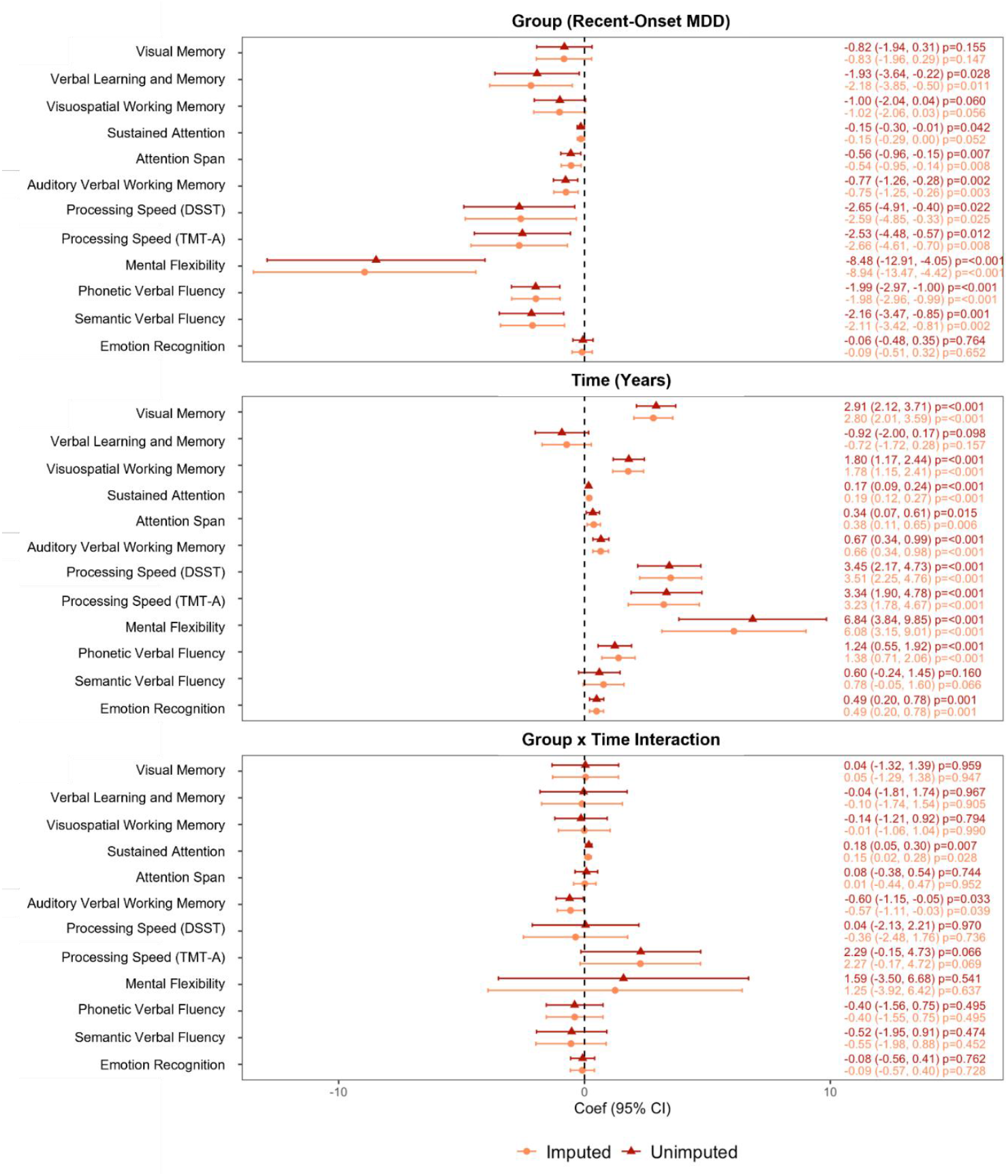
Forest plot showing parameter estimates for fixed effects from linear mixed effects models controlling for age and sex *Reverse-scored so that higher scores indicate better performance.

**Figure 2.**
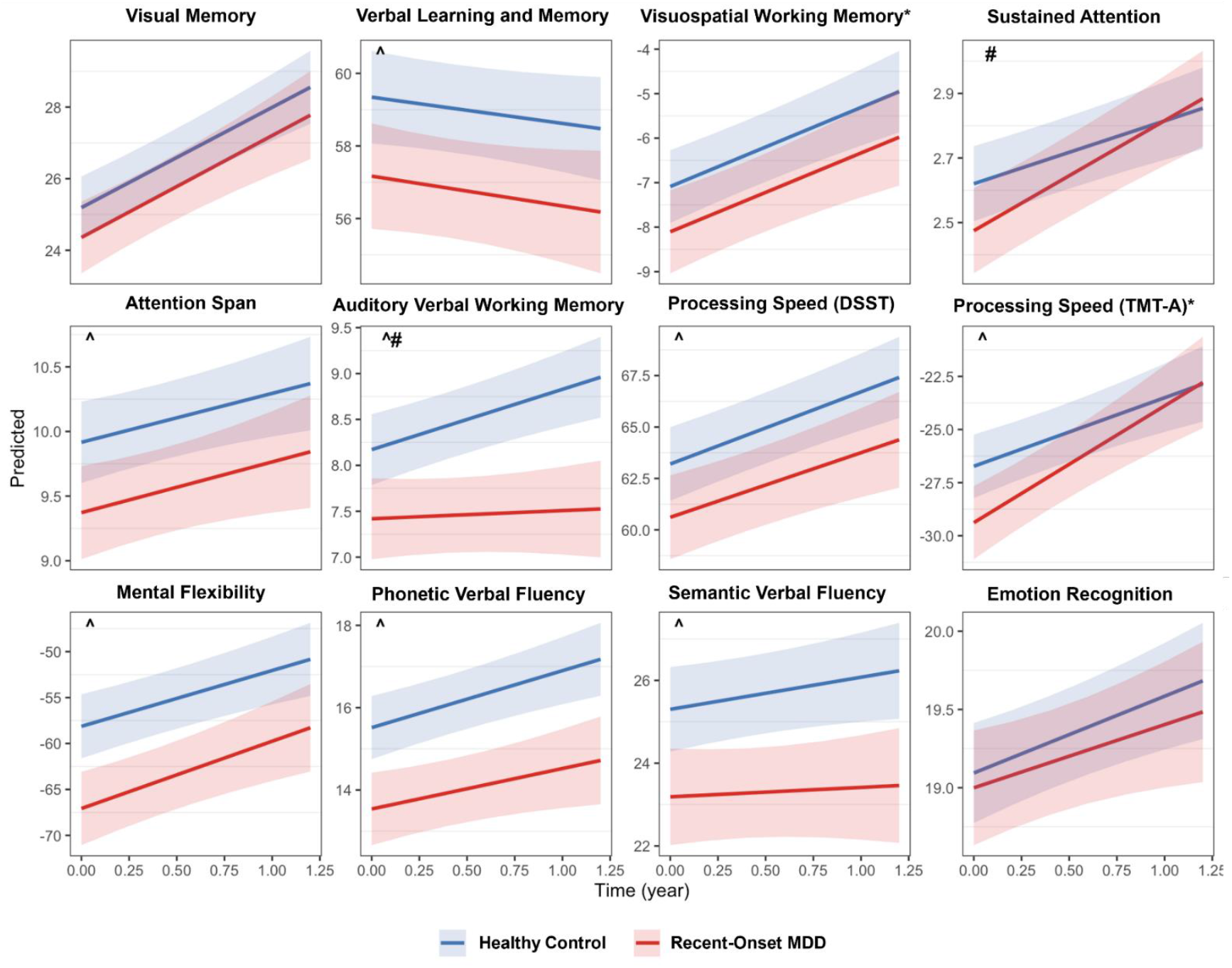
Predicted Cross-level interaction (+/- standard error) showing the Relationship between Time and Cognitive Performance (Cognitive Trajectory) for the Recent-Onset MDD and Healthy Control Groups *Reverse-scored so that higher scores indicate better performance. ^Significant variation in intercept. ^#^Significant variation in slope.

### Cognitive Trajectories

#### No Deficit

As shown in Figures 1 and 2, the ROD group showed no strong evidence suggesting deficit relative to HCs on tests of visual memory, visuospatial working memory, and emotion recognition, with the average performance of both groups significantly improving over time. However, the estimated scores for all these outcomes were lower in the ROD group compared with HCs.

#### Stable Deficit

Though the performance of both groups improved significantly over time, the ROD group showed stable deficits relative to HC on tests of verbal learning and memory, attention, processing speed, mental flexibility, phonetic and semantic verbal fluency.

#### Lag

A lag trajectory in auditory verbal working memory was observed for the ROD group, with a significant group-by-time interaction. While the HC made significant improvements over time, performance of the ROD group remained relatively stable, with increasing divergence between the two groups over time.

#### Deterioration

No absolute deterioration in test performance was observed in either group over the nine-month follow-up period.

#### Catch-Up

Sustained attention demonstrated a catch-up trajectory for the ROD group. A significant group-by-time interaction showed that the ROD group made a greater improvement over time than the HCs, with baseline deficits no longer present at follow-up.

### Relationship to Depressive Symptoms

Most cognitive variables were found to be associated with baseline differences in depression symptoms (between-person variations) except for sustained attention and emotion recognition (Figure 3). Among the cognitive variables with between-person associations, most also demonstrated within-person associations, indicating that changes in cognitive outcomes were linked to changes in depressive symptoms over time, with the exceptions of verbal learning and memory and phonetic and semantic fluency.

**Figure 3.**
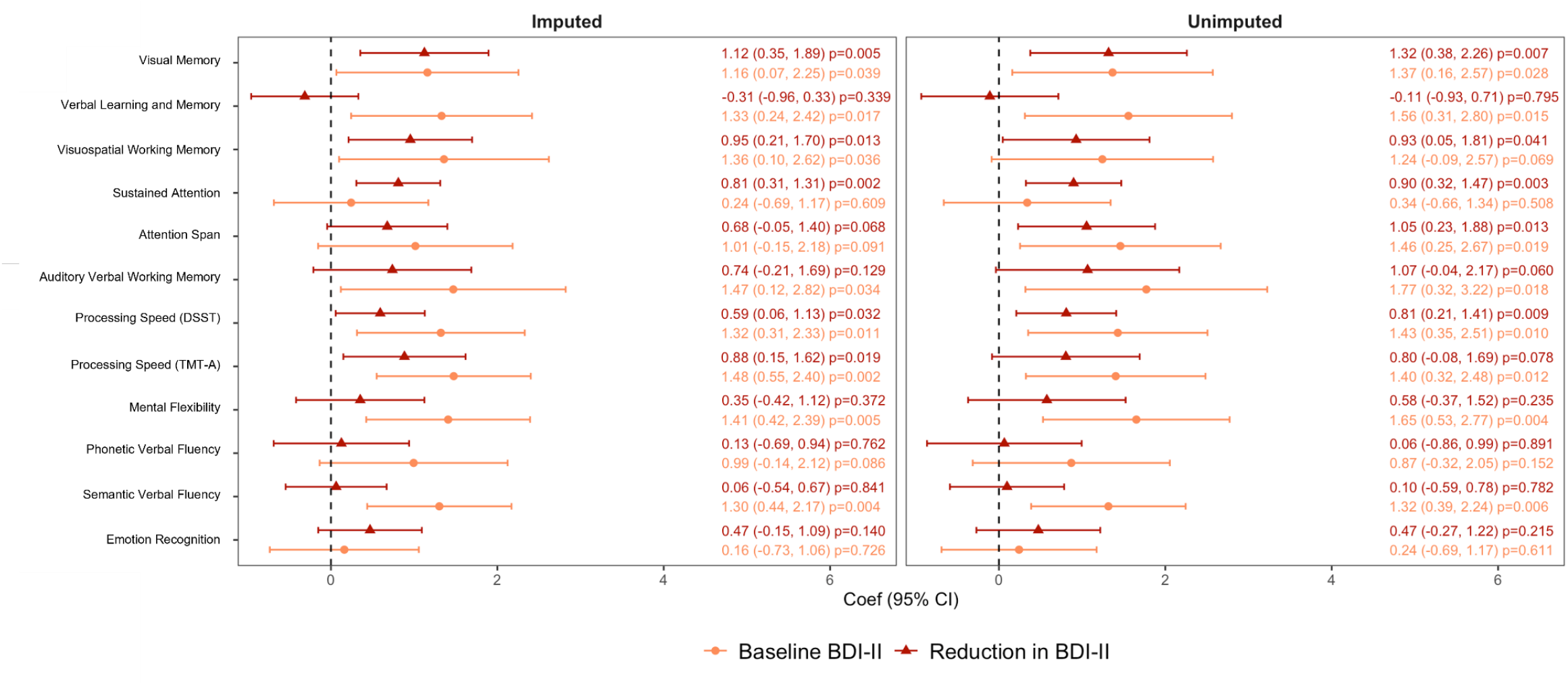
Baseline-centred change in cognitive test scores in relation to reduction in depressive symptomatology in the Recent-Onset MDD group. *Reverse-scored so that higher scores indicate better performance

Improvements on the sustained attention task showed a strong and significant association with reductions in depressive symptoms over time, despite the absence of between-person associations.

When the associations between sustained attention performance and depressive symptoms were plotted (Supplementary Figure 2), the results suggest that even mild depressive symptoms are linked to significant impairments in sustained attention. However, this relationship appears to plateau as depressive symptoms increase, which may explain the absence of between-person associations.

Sensitivity analyses conducted using the same models fitted with GAM confirmed an identical pattern of findings and thus were not reported here. Sensitivity analyses scaled to minimum-maximum cognitive test scores (Supplementary Figures 3 and 4), as well as and excluding participants with multiple MDD episodes (Supplementary Figures 5 and 6), also confirmed the same pattern of cognitive trajectories over time.

## Discussion

We investigated the longitudinal trajectories of cognitive performance in the nine months following ROD. To our knowledge, this is the largest study to date with a well-matched HC group also tested at two timepoints, using a comprehensive cognitive assessment. Performance of the overall study population improved for all cognitive functions examined, except for verbal learning and memory, and semantic verbal fluency, which remained stable. Cognitive domains followed four distinct trajectories following ROD (*deficit, lag, catch-up* and *no-deficit*). While improvements in cognitive performance were associated with reductions in depressive symptom scores over the follow-up period, some deficits relative to HC remained or even increased. As such, this study lends weight to the idea that treatments targeting cognition (e.g., remediation) should be initiated alongside standard depression treatments from the first episode of MDD and tailored to the individual’s unique profile of strengths and weaknesses, as well as their preferences.

The ROD group demonstrated a stable deficit on tests of verbal learning and memory, attention, processing speed, mental flexibility, phonetic verbal fluency, and semantic verbal fluency. Another group also observed enduring executive function (including switching tasks and semantic verbal fluency) and processing speed deficits up to five years following first-episode MDD (10, 12). Here, change in performance on verbal learning and memory, phonetic and semantic verbal fluency was unrelated to reductions in depressive symptomatology over the follow-up period, possibly indicating that these verbally mediated cognitive domains may reflect trait markers of illness (5).

Indeed, poorer verbal learning and memory has been associated with increased risk for depression onset (35) and verbal/language impairments have been observed in unaffected first-degree relatives, suggesting a genetic role in cognitive impairment in MDD (36).

A lag in auditory verbal working memory was observed in the ROD group, whose average test performance remained stable, while HC improved. This is consistent with the findings of a previous study (37). In our analyses, change in auditory verbal working memory performance was related to reductions in depressive symptomatology, but with a smaller effect size than the baseline variation. Together, these findings suggest that the onset of MDD during adolescence/young adulthood may be associated with ‘developmental arrest’ in working memory functioning, which typically continues to develop into the thirties (9). Longer-term follow-up is needed in future studies with younger samples to further test this hypothesis. It also cannot be ruled out that this could represent an absence of practice effects in the ROD group on this test.

Sustained attention performance was poorer on average in the ROD group than the HC group, but the rate of improvement was significantly greater, demonstrating a catch-up trajectory. Although a previous meta-analysis has demonstrated impaired sustained attention during the first episode of depression (38), less is known about its longitudinal course. A meta-analysis of cross-sectional data suggests a small, enduring, impairment in visual selective attention (39), while one of the only other longitudinal studies of ROD patients demonstrated a significant improvement in attention over a six-month period (11). However, their analysis was limited to individuals who remitted from their depressive episode following treatment (11). The present analyses showed a strong relationship between reduction in depressive symptoms over time and improvement in test performance, suggestive of a potential state marker, consistent with the ‘trouble concentrating’ symptom of MDD and in alignment with the previous study of remitted patients (11).

Visual memory, visuospatial working memory, and emotion recognition were the only unimpaired cognitive functions over time in the ROD group. This finding is somewhat inconsistent with cross-sectional meta-analyses (2, 39), and could require a larger sample size to detect small effects. However, it does suggest that some cognitive functions may be spared at illness onset.

## Limitations

Sample size was insufficient to reliably investigate age-related differences in cognitive trajectories. Differences in normative cognitive trajectory might be expected within the range of included participants (15 to 40 years), given performance of many cognitive abilities peaks before age 30 (9). Consistent with this, when other variables were held constant, age was a significant predictor of several cognitive functions (Supplementary Table 5). There may be a ceiling effect within our analysis of the relationship of cognitive trajectories to clinical course, given that this was a moderately depressed sample at baseline which generally improved over time and only 10% experienced a second depressive episode (Table 1 and Supplementary Table 3). However, ongoing depressive symptoms were still evident on average at follow-up. We also did not have a measurement of cognitive performance prior to the onset of ROD. Thus, we cannot rule out the possibility that purported trait impairments are ‘scars’ from a first depressive episode (or a combination of both). This analysis was based on two time points, which may limit the generalisability and robustness of the findings. Finally, treatment (pharmaceutical or psychological), or the absence of treatment, were not accounted for in this analysis. These uncontrolled aspects of the study design may have accounted for some of the unexplained variance in the MLMs.

## Implications

Future work should build on the relative performance trajectory approach that has been utilised in this study, including important clinical and demographic variables such as age and treatment modality. This approach is already established in the psychosis literature (40). Consistency between fields may be useful as clinical focus shifts towards a transdiagnostic approach (41). The association between cognitive impairments and functional outcomes in ROD should also be investigated (41). Longer-term follow-up will also help to track cognition and clinical course in this early-stage group who may also experience diagnostic change over time.

Overall, the findings from this study demonstrate differential cognitive trajectories in the nine months following ROD, irrespective of clinical course. Understanding how cognitive performance changes over time with MDD onset and progression will have important treatment and prognostic implications, especially given the dynamic relationship between cognition and social, familial, inter-personal, and vocational functioning (3, 4). Young people with mental illness report that cognitive impairment is distressing, disabling and want effective treatments (42). Therefore, knowledge of differential cognitive trajectories may inform tailored intervention approaches seeking to improve deficits, prevent further deterioration, and harness cognitive strengths to preserve intact functions in accordance with the wishes of the individual.

## PRONIA Consortium members

Nikolaos Koutsouleris, Dominic B. Dwyer, Lana Kambeitz-Ilankovic, Anne Ruef, Alkomiet Hasan, Claudius Hoff, Ifrah Khanyaree, Aylin Melo, Susanna Muckenhuber-Sternbauer, Yanis Köhler, Ömer Öztürk, Nora Penzel, David Popovic, Adrian Rangnick, Sebastian von Saldern, Rachele Sanfelici, Moritz Spangemacher, Ana Tupac, Maria Fernanda Urquijo, Johanna Weiske, Antonia Wosgien, Camilla Krämer, Shalaila S. Haas, Rebekka Lencer, Inga Meyhoefer, Marian Surmann, Udo Dannlowski, Olga Bienek, Georg Romer, Marlene Rosen, Theresa Lichtenstein, Stephan Ruhrmann, Joseph Kambeitz, Karsten Blume, Dominika Julkowski, Nathalie Kaden, Ruth Milz, Alexandra Nikolaides, Mauro Silke Vent, Martina Wassen, Christos Pantelis, Stefan Borgwardt, Christina Andreou, André Schmidt, Anita Riecher-Rössler, Laura Egloff, Fabienne Harrisberger, Ulrike Heitz, Claudia Lenz, Letizia Leanza, Amatya Mackintosh, Renata Smieskova, Erich Studerus, Anna Walter, Sonja Widmayer, Alexandra Korda, Rachel Upthegrove, Chris Day, Sian Lowri Griffiths, Mariam Iqbal, Mirabel Pelton, Pavan Mallikarjun, Alexandra Stainton, Ashleigh Lin, Paris Lalousis, Raimo K. R. Salokangas, Alexander Denissoff, Anu Ellilä, Tiina From, Markus Heinimaa, Tuula Ilonen, Päivi Jalo, Heikki Laurikainen, Antti Luutonen, Akseli Mäkela, Janina Paju, Henri Pesonen, Reetta-Liina Säilä, Anna Toivonen, Otto Turtonen, Frauke Schultze-Lutter, Eva Meisenzahl, Sonja Botterweck, Norman Kluthausen, Gerald Antoch, Julian Caspers, Hans-Jörg Wittsack, Ana Beatriz Solana, Manuela Abraham, Timo Schirmer, Alessandro Bertolino, Linda A. Antonucci, Giulio Pergola, Ileana Andriola, Barbara Gelao, Paolo Brambilla, Carlo Altamura, Marika Belleri, Francesca Bottinelli, Adele Ferro, Marta Re, Emiliano Monzani, Maurizio Sberna, Armando D’Agostino, Lorenzo Del Fabro, Giampaolo Perna, Maria Nobile, Alessandra Alciati, Matteo Balestrieri, Carolina Bonivento, Giuseppe Cabras, Franco Fabbro, Marco Garzitto & Sara Piccin

## Data Availability

Data are not available for this manuscript due to agreements confirmed at the start of the study.

## Acknowledgments

The authors would also like to acknowledge the significant contributions of the PRONIA consortium members who performed the screening, recruitment, rating, examination, and follow-up of the study participants. They were involved in implementing the examination protocols of the study, setting up its IT infrastructure, and organizing the flow and quality control of the data analysed in this manuscript between the local study sites and the central study database.

