## Supplementary for "Cognitive Trajectories in the Nine Months following Recent-Onset Major Depressive Disorder"

### Methods

**Supplementary Table 1.** *Inclusion and Exclusion Criteria for the Present Study*
**Supplementary Figure 1.** *Overview of PRONIA Dataset Sample Selected for Inclusion in the Current Study*
**Supplementary Table 2.** *Cognitive Testing Battery, Cognitive Function and Performance Measures used in the Current Study*

### Results

**Supplementary Table 3.** *Clinical Characteristics of the Recent-Onset Major Depression Disorder Group*
**Supplementary Table 4.** *Mean, Standard Deviation and Range of Cognitive Test Performance Variables*
**Supplementary Figure 2.** *Scatter Plots with Locally Estimated Scatterplot Smoothing for Each Outcome Variable Against BDI-II Scores at Baseline in the Recent-Onset MDD Group*
**Supplementary Table 5.** *Linear mixed effects regression for each cognitive test*
**Supplementary Figure 3.** *Sensitivity Analyses Scaled to the Minimum-Maximum Test Scores. Forest plot showing parameter estimates for fixed effects.*
**Supplementary Figure 4.** *Sensitivity Analyses Scaled to the Minimum-Maximum Test Scores. Predicted Cross-level interaction (+/- standard error) showing the Relationship between Time and Cognitive Performance (Cognitive Trajectory) for the Recent-Onset MDD and Healthy Control Groups*
**Supplementary Figure 5.** *Sensitivity Analyses in Single-Episode Subset of Recent-Onset MDD Group. Forest plot showing parameter estimates for fixed effects.*
**Supplementary Figure 6.** *Sensitivity Analyses in Single-Episode Subset of Recent-Onset MDD Group. Predicted Cross-level interaction (+/- standard error) showing the Relationship between Time and Cognitive Performance (Cognitive Trajectory) for the Recent-Onset MDD and Healthy Control Groups*

### Methods

**Supplementary Table 2.** *Inclusion and Exclusion Criteria for the Present Study*

| | Criteria | |
|---|---|---|
| | Inclusion | Exclusion |
| General | ▪ Aged 15 and 40 years at enrolment<br>▪ capacity to provide informed consent, or assent for participants <18 years<br>▪ sufficient language ability for study participation | ▪ IQ <70<br>▪ current or past head trauma with loss of consciousness (>5 minutes)<br>▪ neurological or somatic disorder with potential to affect brain function and/or structure<br>▪ alcohol dependence<br>▪ polysubstance dependence within the preceding 6 months<br>▪ medical contraindication for Magnetic Resonance Imaging<br>▪ no cognitive test results<br>▪ follow-up duration unknown or outside the target range (9 +/- 3 months). |
| Recent-Onset MDD group | ▪ fulfilment of DSM-IV-TR major depression criteria within the preceding three months as determined by Structural Clinical Interview for DSM-IV-TR Research Version for Patients(1) | ▪ a previous DSM-IV-TR major depressive episode<br>▪ chronicity (duration >24 months) of the recent/current episode<br>▪ intake of antipsychotic medication (>30 cumulative days) or drugs (within three months preceding baseline) at or above a specified minimum dosage threshold(2)<br>▪ DSM-IV-TR major depressive episode with psychotic features, or with primary bipolar disorder diagnosis at follow-up.<br>▪ Anomalous data (e.g., no MDD episodes). |
| Healthy control group | | ▪ current or past DSM-IV axis I disorder<br>▪ Meeting criteria for a Clinical High Risk for psychosis state<br>▪ affective or non-affective psychosis in a first-degree relative<br>▪ psychotropic medication/drug intake >5 times/year and in the month preceding enrolment. |

**Supplementary Figure 1.** *Overview of PRONIA Dataset Sample Selected for Inclusion in the Current Study*

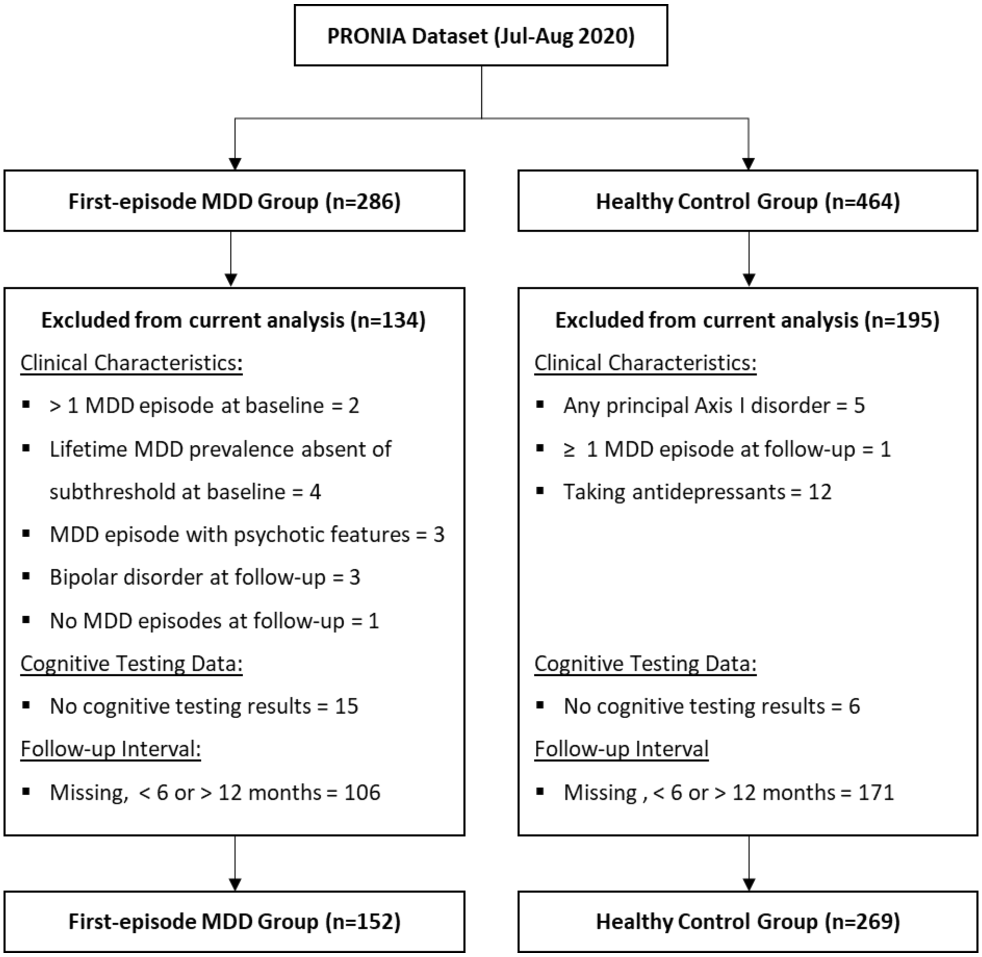

**Supplementary Table 2.** *Cognitive Testing Battery, Cognitive Function and Performance Measures used in the Current Study*

| Cognitive test | Abbreviation | Cognitive function measured | Description | Score range | |
|---|---|---|---|---|---|
| Rey-Osterrieth complex figure test (3, 4) | ROCF | Visual memory | Immediate recall score | 0 | 36 |
| Rey auditory verbal learning test (5) | RAVLT | Verbal learning and memory | Total recall (trials 1 to 5) | 0 | 75 |
| Self-ordered pointing task (6) | SOPT | Visuospatial working memory | Number of errors | 0 | 84 |
| Continuous performance test (7) | CPT | Sustained attention | Discriminability (d')[a] | 0 | 5 |
| Digit-span – forward (8) | DSF | Attention span | Number of correct trials | 0 | 16 |
| Digit-span – backward (8) | DSB | Auditory verbal working memory | Number of correct trials | 0 | 16 |
| Digit-symbol substitution test (9) | DSST | Processing speed | Digit-symbol match score | 0 | 120 |
| Trail making test – A (10) | TMT-A | Processing speed | Execution time (seconds)[b] | 5 | 300 |
| Trail making test – B (10) | TMT-B | Mental flexibility | Execution time (seconds)[b] | 5 | 300 |
| Phonetic verbal fluency (11) | PVF | Phonetic verbal fluency | Number of correct unique words | 0 | unlimited |
| Semantic verbal fluency (11) | SVF | Semantic verbal fluency | Number of correct unique words | 0 | unlimited |
| Diagnostic analysis of nonverbal accuracy-II (12) | DANVA2 | Emotion recognition | Number of faces correct | 0 | 24 |

*Note.* At both time-points the test battery was identical and administered in a standardised order.
[a] Discriminability is an unbounded statistic; a d' of zero indicates performance at chance level (i.e., guessing) and a higher d' is indicative of better discriminability
[b] Execution times that were negative (impossible), less than five seconds or greater than five minutes were excluded from analysis.

### Results

**Supplementary Table 3.** *Clinical Characteristics of the Recent-Onset Major Depression Disorder Group*

| | Baseline | | | Follow-up | | |
|---|---|---|---|---|---|---|
| | N | *n* | % | N | *n* | % |
| **Antidepressants** | 152 | 118 | 77.6 | 152 | 98 | 64.5 |
| **Current episode severity** | | | | | | |
| Mild | 138 | 18 | 13 | 78 | 27 | 35 |
| Moderate | 138 | 63 | 46 | 78 | 35 | 45 |
| Severe | 138 | 57 | 41 | 78 | 16 | 21 |
| **Lifetime MDD episodes** | | | | | | |
| 1 | 148 | 148 | 100 | 132 | 119 | 90 |
| >1 | 148 | 0 | 0 | 132 | 13 | 10 |

*Note.* Current depression severity and number of episodes was determined through the Structural Clinical Interview for DSM-IV-TR Research Version for Patients (SCID-I/P) conducted at both time-points

**Supplementary Table 4.** *Mean, Standard Deviation and Range of Cognitive Test Performance Variables*

| **Cognitive test Performance variable** | Recent-Onset MDD | | | | | Healthy control | | | | |
|---|---|---|---|---|---|---|---|---|---|---|
| | N | *M* | *SD* | Range | | N | *M* | *SD* | Range | |
| | | | | Min | Max | | | | Min | Max |
| **ROCF**, Immediate recall score | | | | | | | | | | |
| Baseline (T0) | 148 | 24.2 | 6.4 | 2.0 | 36.0 | 268 | 25 | 5.5 | 8 | 36 |
| Follow-up (T1) | 135 | 26.6 | 6.1 | 11.0 | 36.0 | 250 | 27.3 | 5 | 14 | 36 |
| Change (T1 - T0) | 131 | 2.1 | 5.4 | -12.5 | 16.5 | 249 | 2.3 | 5.2 | -16 | 18.5 |
| **RAVLT**, Number of correct trials | | | | | | | | | | |
| Baseline (T0) | 136 | 58.4 | 7.9 | 31.0 | 75.0 | 226 | 60.6 | 7.6 | 40 | 73 |
| Follow-up (T1) | 127 | 57.7 | 8.6 | 31.0 | 75.0 | 209 | 60 | 8.6 | 29 | 75 |
| Change (T1 - T0) | 123 | -0.9 | 6.3 | -23 | 14 | 207 | -0.9 | 6.7 | -27 | 14 |
| **Self-ordered pointing task**, Number of errors | | | | | | | | | | |
| Baseline (T0) | 147 | -8.6 | 5.9 | -27 | 0 | 264 | -7.6 | 5 | -21 | 0 |
| Follow-up (T1) | 138 | -7.1 | 5.6 | -28 | 0 | 249 | -6.1 | 4.7 | -26 | 0 |
| Change (T1 - T0) | 133 | 1.3 | 4.2 | -12 | 16 | 245 | 1.5 | 4.2 | -14 | 11 |
| **Continuous performance**, Discriminability (d’) | | | | | | | | | | |
| Baseline (T0) | 147 | 2.4 | 0.8 | -0.1 | 4.5 | 266 | 2.5 | 0.7 | 0.5 | 4.3 |
| Follow-up (T1) | 137 | 2.7 | 0.9 | 0.7 | 5.3 | 250 | 2.7 | 0.7 | 1.1 | 4.9 |
| Change (T1 - T0) | 132 | 0.3 | 0.5 | -0.9 | 1.9 | 247 | 0.2 | 0.5 | -1.1 | 2.2 |
| **Digit span – forward**, Number of correct trials | | | | | | | | | | |
| Baseline (T0) | 147 | 9.3 | 2.0 | 4.0 | 14.0 | 266 | 9.8 | 2 | 0 | 15 |
| Follow-up (T1) | 138 | 9.6 | 2.1 | 4.0 | 15.0 | 251 | 10.1 | 2 | 5 | 15 |
| Change (T1 - T0) | 133 | 0.4 | 1.6 | -4 | 6 | 249 | 0.3 | 1.9 | -5 | 7 |
| **Digit span – back**, Number of correct trials | | | | | | | | | | |
| Baseline (T0) | 147 | 7.4 | 2.3 | 3.0 | 13.0 | 267 | 8.1 | 2.4 | 0 | 14 |
| Follow-up (T1) | 137 | 7.5 | 2.7 | 2.0 | 13.0 | 251 | 8.6 | 2.4 | 2 | 14 |
| Change (T1 - T0) | 132 | 0.1 | 2.3 | -6 | 6 | 249 | 0.6 | 2.1 | -6 | 6 |
| **Digit-symbol substitution**, Digit-symbol match score | | | | | | | | | | |
| Baseline (T0) | 148 | 62.1 | 11.8 | 30.0 | 91.0 | 268 | 65.1 | 11.4 | 0 | 105 |
| Follow-up (T1) | 137 | 64.9 | 12.4 | 35.0 | 100.0 | 251 | 68 | 10.4 | 44 | 103 |
| Change (T1 - T0) | 133 | 2.8 | 7.4 | -15 | 28 | 250 | 2.8 | 9.1 | -26 | 73 |
| **Trail making test – A**, Execution time | | | | | | | | | | |
| Baseline (T0) | 141 | -29.7 | 12.5 | -78.1 | -6.6 | 260 | -27.1 | 8.9 | -63 | -12 |
| Follow-up (T1) | 130 | -24.9 | 9.7 | -74.4 | -11.3 | 233 | -24.2 | 8 | -61.1 | -8.6 |

| Cognitive test<br>Performance variable | Recent-Onset MDD | | | | | Healthy control | | | | |
|---|---|---|---|---|---|---|---|---|---|---|
| | N | *M* | *SD* | Range | | N | *M* | *SD* | Range | |
| | | | | Min | Max | | | | Min | Max |
| Change (T1 - T0) | 119 | 4.2 | 10.2 | -24.4 | 52.2 | 224 | 2.4 | 8.5 | -29.1 | 29.9 |
| **Trail making test – B**, Execution time | | | | | | | | | | |
| Baseline (T0) | 140 | -65.4 | 27.8 | -186 | -26.6 | 259 | -56 | 18.5 | -165.3 | -22.2 |
| Follow-up (T1) | 130 | -58.5 | 26.1 | -177.7 | -26.8 | 233 | -50.5 | 17.9 | -155 | -20.4 |
| Change (T1 - T0) | 118 | 6.7 | 26.1 | -121.1 | 129 | 223 | 5.5 | 14 | -53.6 | 66.1 |
| **Phonetic Verbal Fluency**, Number of words | | | | | | | | | | |
| Baseline (T0) | 147 | 14 | 4.6 | 3 | 27 | 267 | 16 | 5.1 | 0 | 31 |
| Follow-up (T1) | 137 | 14.6 | 5.2 | 3 | 29 | 250 | 17.2 | 4.9 | 4 | 31 |
| Change (T1 - T0) | 132 | 0.8 | 4.5 | -18 | 16 | 248 | 1.1 | 4.6 | -12 | 26 |
| **Semantic verbal fluency**, Number of words | | | | | | | | | | |
| Baseline (T0) | 147 | 23.4 | 6.6 | 4 | 39 | 267 | 25.6 | 6 | 10 | 45 |
| Follow-up (T1) | 134 | 23.6 | 7.2 | 4 | 49 | 249 | 26.3 | 6.7 | 9 | 61 |
| Change (T1 - T0) | 129 | 0.2 | 5.6 | -14 | 32 | 247 | 0.6 | 5.5 | -14 | 38 |
| **DANVA-2**, Number of faces correct | | | | | | | | | | |
| Baseline (T0) | 147 | 19.3 | 2.1 | 11 | 23 | 267 | 19.4 | 2.1 | 13 | 24 |
| Follow-up (T1) | 138 | 19.6 | 2.1 | 9 | 23 | 251 | 19.8 | 2 | 12 | 23 |
| Change (T1 - T0) | 133 | 0.4 | 1.9 | -5 | 5 | 249 | 0.4 | 2 | -5 | 8 |

*Note.* DANVA = Diagnostic analysis of nonverbal accuracy-2; MDD = Major depressive disorder; RAVLT = Rey auditory verbal learning task; ROCF = Rey-Osterrieth complex figure test.

**Supplementary Figure 2.** *Scatter Plots with Locally Estimated Scatterplot Smoothing for Each Outcome Variable Against BDI-II Scores at Baseline in the Recent-Onset MDD Group*

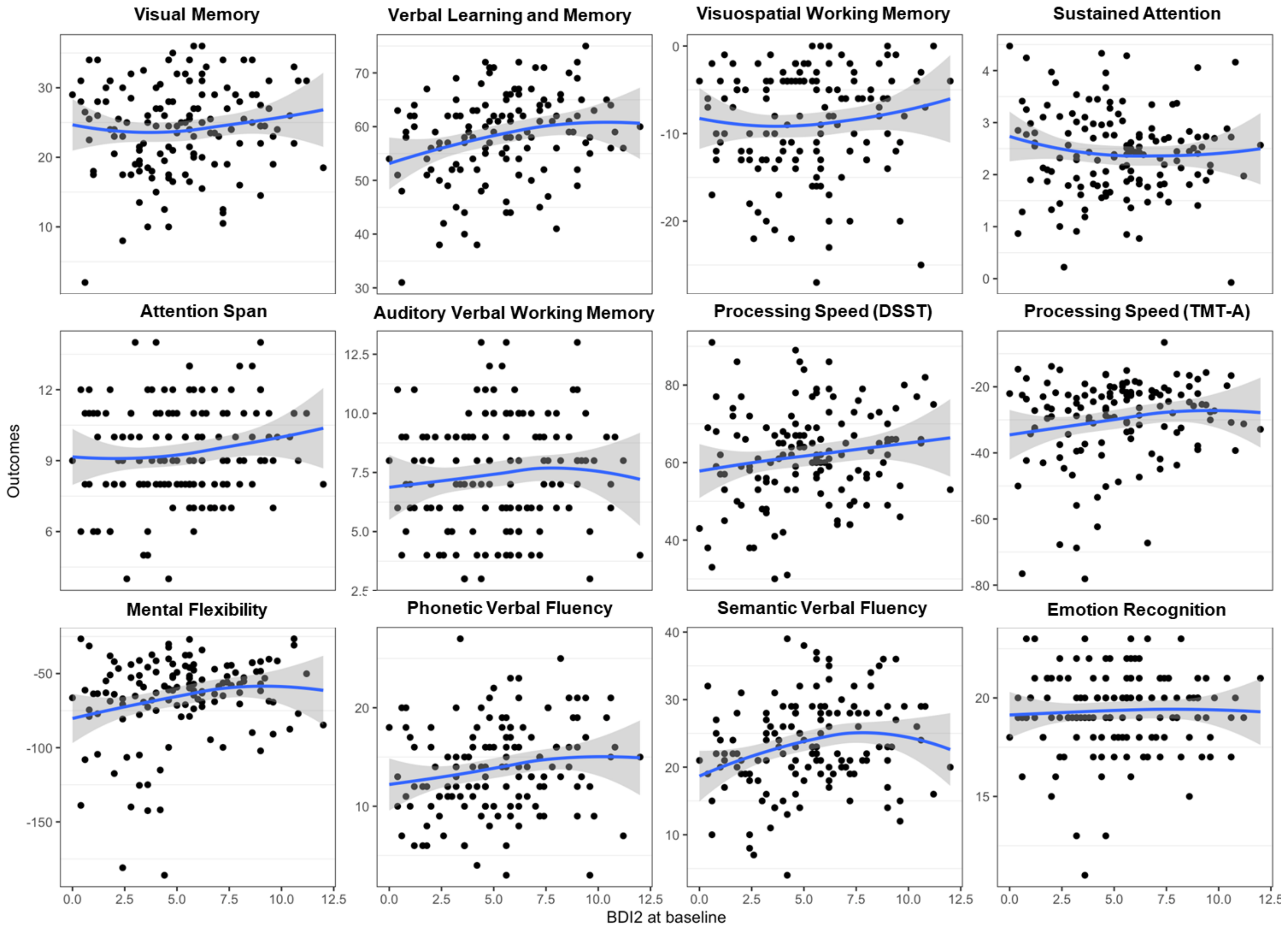

**Supplementary Table 5.** *Linear mixed effects regression for each cognitive test*

| **Rey-Osterrieth Complex Figure (ROCF)** | | | | |
|---|---|---|---|---|
| | **Unimputed** | | **Imputed** | |
| **Predictor** | **Coef (95% CI)** | **p-value** | **Coef (95% CI)** | **p-value** |
| Study Group | | | | |
| Healthy Control | Ref | | Ref | |
| Recent-onset MDD | -0.82 (-1.94, 0.31) | 0.155 | -0.73 (-1.86, 0.39) | 0.202 |
| Time (Years) | 2.91 (2.12, 3.71) | <0.001 | 2.78 (1.99, 3.56) | <0.001 |
| Group*Time | 0.04 (-1.32, 1.39) | 0.959 | 0.12 (-1.20, 1.43) | 0.862 |
| Age (Years) | -0.12 (-0.20, -0.04) | 0.004 | -0.11 (-0.19, -0.03) | 0.006 |
| Sex | | | | |
| Males | Ref | | Ref | |
| Females | -0.41 (-1.37, 0.56) | 0.410 | -0.26 (-1.21, 0.69) | 0.593 |
| **Random Effects** | **Variance** | **SD** | **Variance** | **SD** |
| Intercept | 17.30 | 4.16 | 17.05 | 4.13 |
| Residual | 14.19 | 3.77 | 14.77 | 3.84 |
| **Rey Auditory Verbal Learning Test (RAVLT)** | | | | |
| | **Unimputed** | | **Imputed** | |
| **Predictor** | **Coef (95% CI)** | **p-value** | **Coef (95% CI)** | **p-value** |
| Study Group | | | | |
| Healthy Control | Ref | | Ref | |
| Recent-onset MDD | -1.93 (-3.64, -0.22) | 0.028 | -2.19 (-3.80, -0.58) | 0.008 |
| Time (Years) | -0.92 (-2.00, 0.17) | 0.098 | -0.50 (-1.41, 0.41) | 0.283 |
| Group x Time | -0.04 (-1.81, 1.74) | 0.967 | -0.45 (-1.98, 1.07) | 0.560 |
| Age (Years) | -0.07 (-0.19, 0.06) | 0.291 | -0.04 (-0.15, 0.08) | 0.551 |
| Sex | | | | |
| Males | Ref | | Ref | |
| Females | 2.59 (1.06, 4.11) | 0.001 | 2.66 (1.23, 4.09) | <0.001 |
| **Random Effects** | **Variance** | **SD** | **Variance** | **SD** |
| Intercept | 43.60 | 6.60 | 45.11 | 6.72 |
| Residual | 21.54 | 4.64 | 19.86 | 4.46 |
| **Self-Ordered Pointing Test (SOPT)** | | | | |
| | **Unimputed** | | **Imputed** | |
| **Predictor** | **Coef (95% CI)** | **p-value** | **Coef (95% CI)** | **p-value** |
| Study Group | | | | |
| Healthy Control | Ref | | Ref | |
| Recent-onset MDD | -1.00 (-2.04, 0.04) | 0.060 | -0.95 (-1.98, 0.08) | 0.072 |
| Time (Years) | 1.80 (1.17, 2.44) | <0.001 | 1.74 (1.11, 2.37) | <0.001 |
| Group*Time | -0.14 (-1.21, 0.92) | 0.794 | -0.18 (-1.23, 0.87) | 0.739 |
| Age (Years) | -0.02 (-0.09, 0.06) | 0.668 | 0.00 (-0.08, 0.07) | 0.958 |
| Sex | | | | |
| Males | Ref | | Ref | |
| Females | -1.03 (-1.95, -0.10) | 0.030 | -0.90 (-1.81, 0.00) | 0.051 |
| **Random Effects** | **Variance** | **SD** | **Variance** | **SD** |
| Intercept | 18.05 | 4.25 | 17.31 | 4.16 |
| Residual | 8.73 | 2.95 | 9.42 | 3.07 |
| **Continuous Performance Test (CPT)** | | | | |
| | **Unimputed** | | **Imputed** | |
| **Predictor** | **Coef (95% CI)** | **p-value** | **Coef (95% CI)** | **p-value** |
| Study Group | | | | |
| Healthy Control | Ref | | Ref | |
| Recent-onset MDD | -0.15 (-0.30, -0.01) | 0.042 | -0.15 (-0.29, 0.00) | 0.051 |
| Time (Years) | 0.17 (0.09, 0.24) | <0.001 | 0.20 (0.13, 0.27) | <0.001 |

| | | | | |
|---|---|---|---|---|
| Group*Time | 0.18 (0.05, 0.30) | 0.007 | 0.15 (0.02, 0.27) | 0.019 |
| Age (Years) | 0.02 (0.01, 0.03) | <0.001 | 0.02 (0.01, 0.03) | <0.001 |
| Sex | | | | |
| Males | Ref | | Ref | |
| Females | -0.14 (-0.27, -0.01) | 0.038 | -0.13 (-0.26, 0.01) | 0.061 |
| **Random Effects** | **Variance** | **SD** | **Variance** | **SD** |
| Intercept | 0.40 | 0.63 | 0.41 | 0.64 |
| Residual | 0.12 | 0.35 | 0.13 | 0.36 |
| **Digit Span Forward (DSF)** | | | | |
| | **Unimputed** | | **Imputed** | |
| **Predictor** | **Coef (95% CI)** | **p-value** | **Coef (95% CI)** | **p-value** |
| Study Group | | | | |
| Healthy Control | Ref | | Ref | |
| Recent-onset MDD | -0.56 (-0.96, -0.15) | 0.007 | -0.49 (-0.89, -0.09) | 0.017 |
| Time (Years) | 0.34 (0.07, 0.61) | 0.015 | 0.31 (0.05, 0.58) | 0.022 |
| Group*Time | 0.08 (-0.38, 0.54) | 0.744 | 0.05 (-0.40, 0.49) | 0.840 |
| Age (Years) | 0.00 (-0.02, 0.03) | 0.780 | 0.01 (-0.02, 0.03) | 0.649 |
| Sex | | | | |
| Males | Ref | | Ref | |
| Females | -0.17 (-0.52, 0.19) | 0.356 | -0.14 (-0.48, 0.21) | 0.429 |
| **Random Effects** | **Variance** | **SD** | **Variance** | **SD** |
| Intercept | 2.42 | 1.55 | 2.34 | 1.53 |
| Residual | 1.64 | 1.28 | 1.72 | 1.31 |
| **Digit Span Backward (DSB)** | | | | |
| | **Unimputed** | | **Imputed** | |
| **Predictor** | **Coef (95% CI)** | **p-value** | **Coef (95% CI)** | **p-value** |
| Study Group | | | | |
| Healthy Control | Ref | | Ref | |
| Recent-onset MDD | -0.77 (-1.26, -0.28) | 0.002 | -0.74 (-1.23, -0.25) | 0.003 |
| Time (Years) | 0.67 (0.34, 0.99) | <0.001 | 0.67 (0.35, 0.98) | <0.001 |
| Group*Time | -0.60 (-1.15, -0.05) | 0.033 | -0.56 (-1.08, -0.04) | 0.036 |
| Age (Years) | 0.01 (-0.03, 0.04) | 0.615 | 0.01 (-0.02, 0.05) | 0.547 |
| Sex | | | | |
| Males | Ref | | Ref | |
| Females | -0.13 (-0.56, 0.30) | 0.555 | -0.07 (-0.49, 0.36) | 0.749 |
| **Random Effects** | **Variance** | **SD** | **Variance** | **SD** |
| Intercept | 3.67 | 1.92 | 3.69 | 1.92 |
| Residual | 2.35 | 1.53 | 2.34 | 1.53 |
| **Digit Symbol Substitution Test (DSST)** | | | | |
| | **Unimputed** | | **Imputed** | |
| **Predictor** | **Coef (95% CI)** | **p-value** | **Coef (95% CI)** | **p-value** |
| Study Group | | | | |
| Healthy Control | Ref | | Ref | |
| Recent-onset MDD | -2.65 (-4.91, -0.40) | 0.022 | -2.54 (-4.80, -0.28) | 0.028 |
| Time (Years) | 3.45 (2.17, 4.73) | <0.001 | 3.43 (2.21, 4.66) | <0.001 |
| Group*Time | 0.04 (-2.13, 2.21) | 0.970 | -0.56 (-2.61, 1.50) | 0.596 |
| Age (Years) | -0.07 (-0.23, 0.10) | 0.419 | -0.07 (-0.23, 0.10) | 0.434 |
| Sex | | | | |
| Males | Ref | | Ref | |
| Females | 3.30 (1.28, 5.32) | 0.001 | 3.35 (1.33, 5.37) | 0.001 |
| **Random Effects** | **Variance** | **SD** | **Variance** | **SD** |
| Intercept | 90.17 | 9.50 | 91.89 | 9.59 |
| Residual | 36.30 | 6.02 | 35.99 | 6.00 |

| **Trail Making Test Part A (TMT-A)** | | | | |
|---|---|---|---|---|
| | **Unimputed** | | **Imputed** | |
| **Predictor** | **Coef (95% CI)** | **p-value** | **Coef (95% CI)** | **p-value** |
| Study Group | | | | |
| Healthy Control | Ref | | Ref | |
| Recent-onset MDD | -2.53 (-4.48, -0.57) | 0.012 | -2.32 (-4.26, -0.38) | 0.019 |
| Time (Years) | 3.34 (1.90, 4.78) | <0.001 | 3.14 (1.76, 4.51) | <0.001 |
| Group*Time | 2.29 (-0.15, 4.73) | 0.066 | 2.04 (-0.26, 4.33) | 0.083 |
| Age (Years) | -0.08 (-0.22, 0.05) | 0.232 | -0.07 (-0.20, 0.07) | 0.332 |
| Sex | | | | |
| Males | Ref | | Ref | |
| Females | -0.91 (-2.57, 0.75) | 0.283 | -0.60 (-2.24, 1.04) | 0.472 |
| **Random Effects** | **Variance** | **SD** | **Variance** | **SD** |
| Intercept | 49.55 | 7.04 | 49.46 | 7.03 |
| Residual | 42.58 | 6.52 | 45.31 | 6.73 |
| **Trail Making Test Part B (TMT-B)** | | | | |
| | **Unimputed** | | **Imputed** | |
| **Predictor** | **Coef (95% CI)** | **p-value** | **Coef (95% CI)** | **p-value** |
| Study Group | | | | |
| Healthy Control | Ref | | Ref | |
| Recent-onset MDD | -8.48 (-12.91, -4.05) | <0.001 | -8.03 (-12.46, -3.60) | <0.001 |
| Time (Years) | 6.84 (3.84, 9.85) | <0.001 | 5.72 (2.98, 8.46) | <0.001 |
| Group*Time | 1.59 (-3.50, 6.68) | 0.541 | 0.47 (-4.10, 5.05) | 0.839 |
| Age (Years) | -0.07 (-0.39, 0.24) | 0.651 | -0.04 (-0.36, 0.28) | 0.814 |
| Sex | | | | |
| Males | Ref | | Ref | |
| Females | 3.52 (-0.32, 7.36) | 0.073 | 4.48 (0.61, 8.35) | 0.024 |
| **Random Effects** | **Variance** | **SD** | **Variance** | **SD** |
| Intercept | 290.06 | 17.03 | 313.53 | 17.71 |
| Residual | 182.78 | 13.52 | 179.22 | 13.39 |
| **Phonetic Verbal Fluency (PVF)** | | | | |
| | **Unimputed** | | **Imputed** | |
| **Predictor** | **Coef (95% CI)** | **p-value** | **Coef (95% CI)** | **p-value** |
| Study Group | | | | |
| Healthy Control | Ref | | Ref | |
| Recent-onset MDD | -1.99 (-2.97, -1.00) | <0.001 | -1.89 (-2.88, -0.90) | <0.001 |
| Time (Years) | 1.24 (0.55, 1.92) | <0.001 | 1.47 (0.81, 2.13) | <0.001 |
| Group*Time | -0.40 (-1.56, 0.75) | 0.495 | -0.44 (-1.55, 0.66) | 0.431 |
| Age (Years) | 0.12 (0.05, 0.19) | 0.001 | 0.13 (0.06, 0.20) | <0.001 |
| Sex | | | | |
| Males | Ref | | Ref | |
| Females | 0.82 (-0.02, 1.66) | 0.057 | 0.89 (0.04, 1.74) | 0.040 |
| **Random Effects** | **Variance** | **SD** | **Variance** | **SD** |
| Intercept | 13.50 | 3.67 | 14.05 | 3.75 |
| Residual | 10.34 | 3.22 | 10.46 | 3.23 |
| **Semantic Verbal Fluency (SVF)** | | | | |
| | **Unimputed** | | **Imputed** | |
| **Predictor** | **Coef (95% CI)** | **p-value** | **Coef (95% CI)** | **p-value** |
| Study Group | | | | |
| Healthy Control | Ref | | Ref | |
| Recent-onset MDD | -2.16 (-3.47, -0.85) | 0.001 | -2.13 (-3.43, -0.83) | 0.001 |
| Time (Years) | 0.60 (-0.24, 1.45) | 0.160 | 0.79 (-0.03, 1.62) | 0.061 |
| Group*Time | -0.52 (-1.95, 0.91) | 0.474 | -0.17 (-1.55, 1.21) | 0.809 |

| | | | | |
|---|---|---|---|---|
| Age (Years) | 0.14 (0.04, 0.23) | 0.004 | 0.14 (0.05, 0.23) | 0.003 |
| Sex | | | | |
| Males | Ref | | Ref | |
| Females | 0.58 (-0.57, 1.73) | 0.323 | 0.59 (-0.54, 1.72) | 0.306 |
| **Random Effects** | **Variance** | **SD** | **Variance** | **SD** |
| Intercept | 26.81 | 5.18 | 26.26 | 5.12 |
| Residual | 15.62 | 3.95 | 16.34 | 4.04 |
| **Diagnostic Analysis of Non-Verbal Accuracy (DANVA2)** | | | | |
| | **Unimputed** | | **Imputed** | |
| **Predictor** | **Coef (95% CI)** | **p-value** | **Coef (95% CI)** | **p-value** |
| Study Group | | | | |
| Healthy Control | Ref | | Ref | |
| Recent-onset MDD | -0.06 (-0.48, 0.35) | 0.764 | -0.07 (-0.47, 0.34) | 0.746 |
| Time (Years) | 0.49 (0.20, 0.78) | 0.001 | 0.51 (0.24, 0.79) | <0.001 |
| Group*Time | -0.08 (-0.56, 0.41) | 0.762 | -0.12 (-0.58, 0.34) | 0.616 |
| Age (Years) | 0.00 (-0.03, 0.02) | 0.755 | 0.00 (-0.03, 0.03) | 0.889 |
| Sex | | | | |
| Males | Ref | | Ref | |
| Females | 0.52 (0.17, 0.87) | 0.004 | 0.53 (0.18, 0.88) | 0.003 |
| **Random Effects** | **Variance** | **SD** | **Variance** | **SD** |
| Intercept | 2.33 | 1.53 | 2.31 | 1.52 |
| Residual | 1.86 | 1.37 | 1.82 | 1.35 |

CI= Confidence Interval

**Supplementary Figure 3.** *Sensitivity Analyses Scaled to the Minimum-Maximum Test Scores. Forest plot showing parameter estimates for fixed effects.*

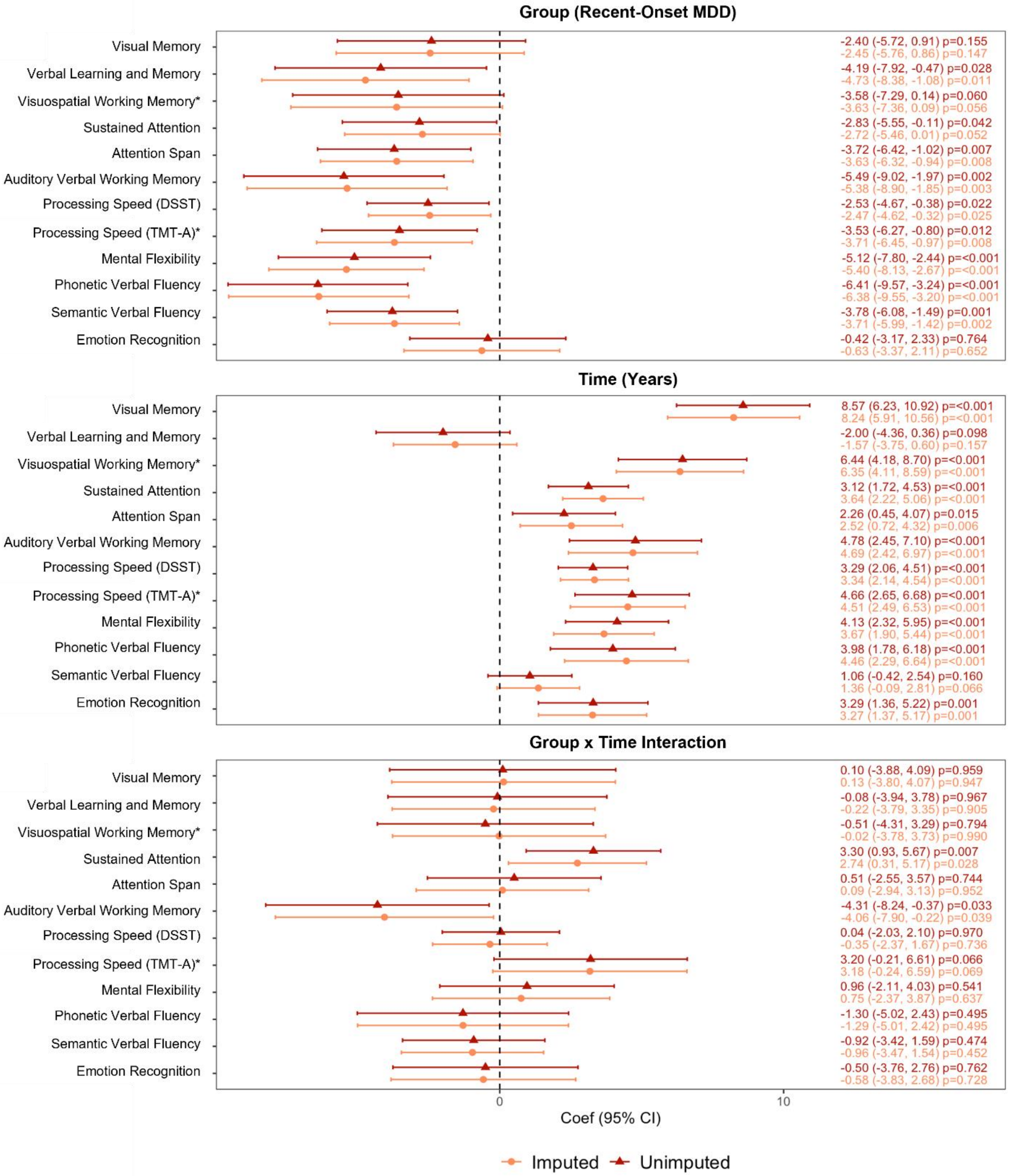


*Reverse-scored so that higher scores indicate better performance.

**Supplementary Figure 4.** *Sensitivity Analyses Scaled to the Minimum-Maximum Test Scores. Predicted Cross-level interaction (+/- standard error) showing the Relationship between Time and Cognitive Performance (Cognitive Trajectory) for the Recent-Onset MDD and Healthy Control Groups*

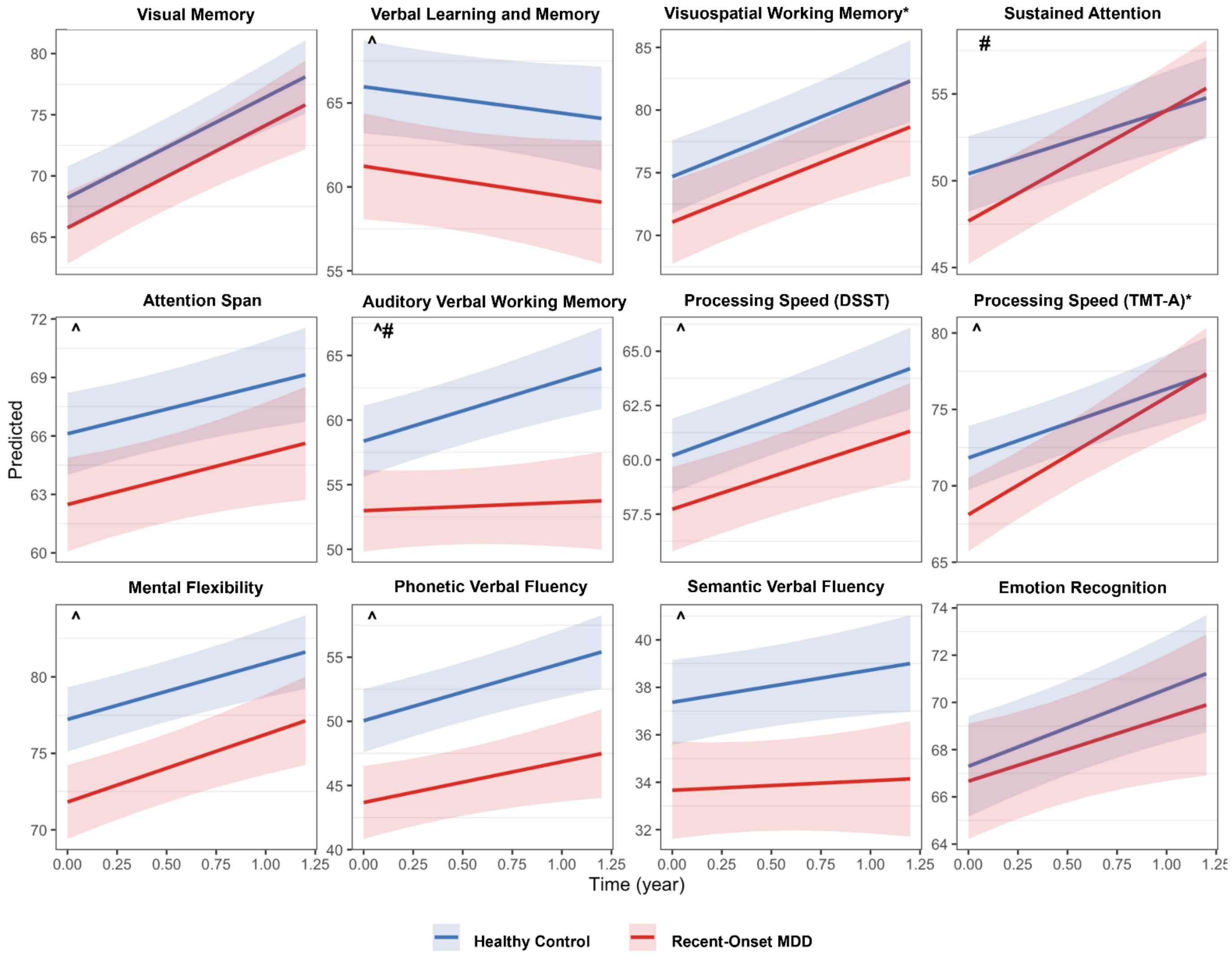


*Reverse-scored so that higher scores indicate better performance. ^Significant variation in intercept. #Significant variation in slope.

**Supplementary Figure 5.** *Sensitivity Analyses in Single-Episode Subset of Recent-Onset MDD Group. Forest plot showing parameter estimates for fixed effects*

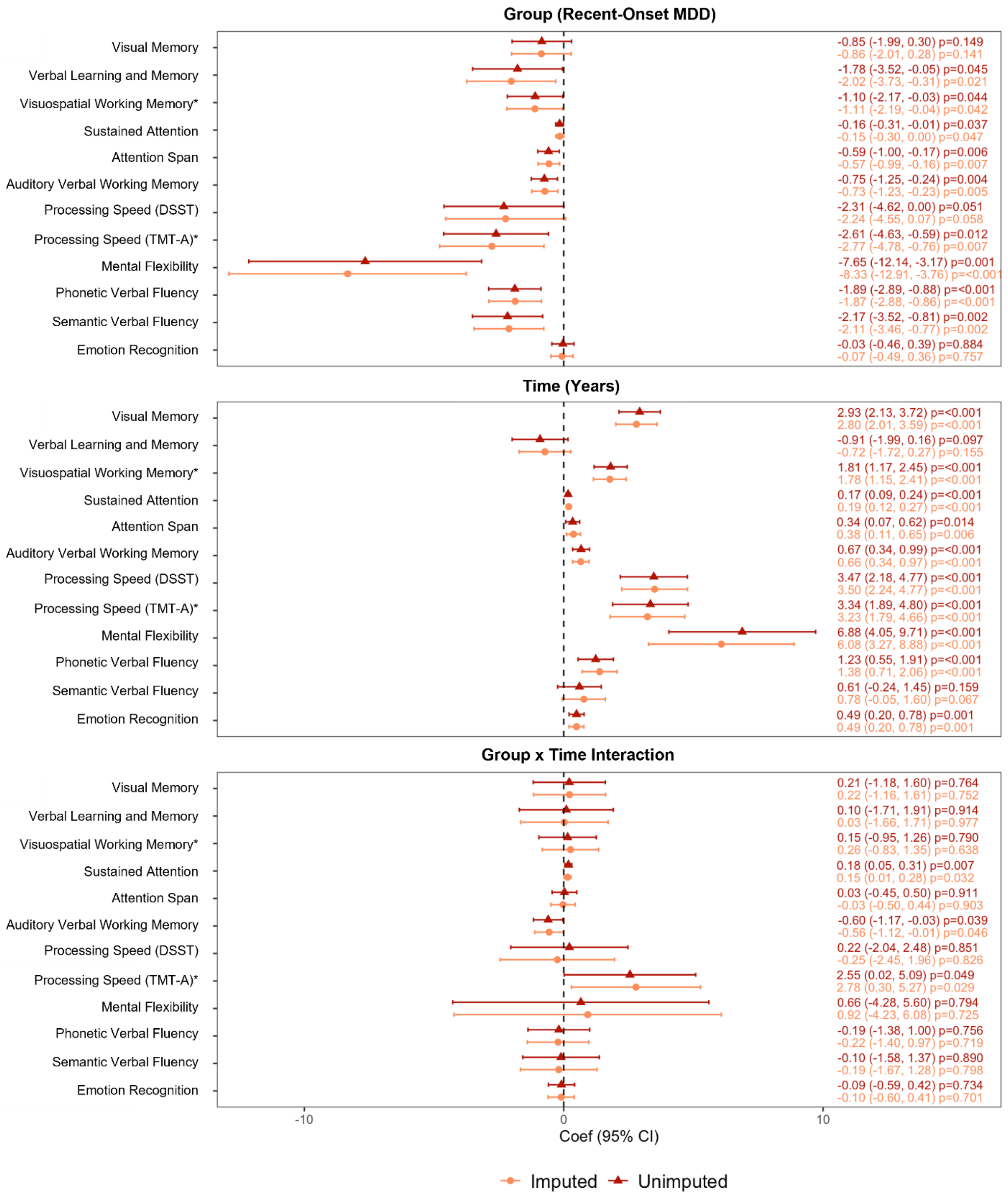


*Reverse-scored so that higher scores indicate better performance.

**Supplementary Figure 6.** *Sensitivity Analyses in Single-Episode Subset of Recent-Onset MDD Group. Predicted Cross-level interaction (+/- standard error) showing the Relationship between Time and Cognitive Performance (Cognitive Trajectory) for the Recent-Onset MDD and Healthy Control Groups*

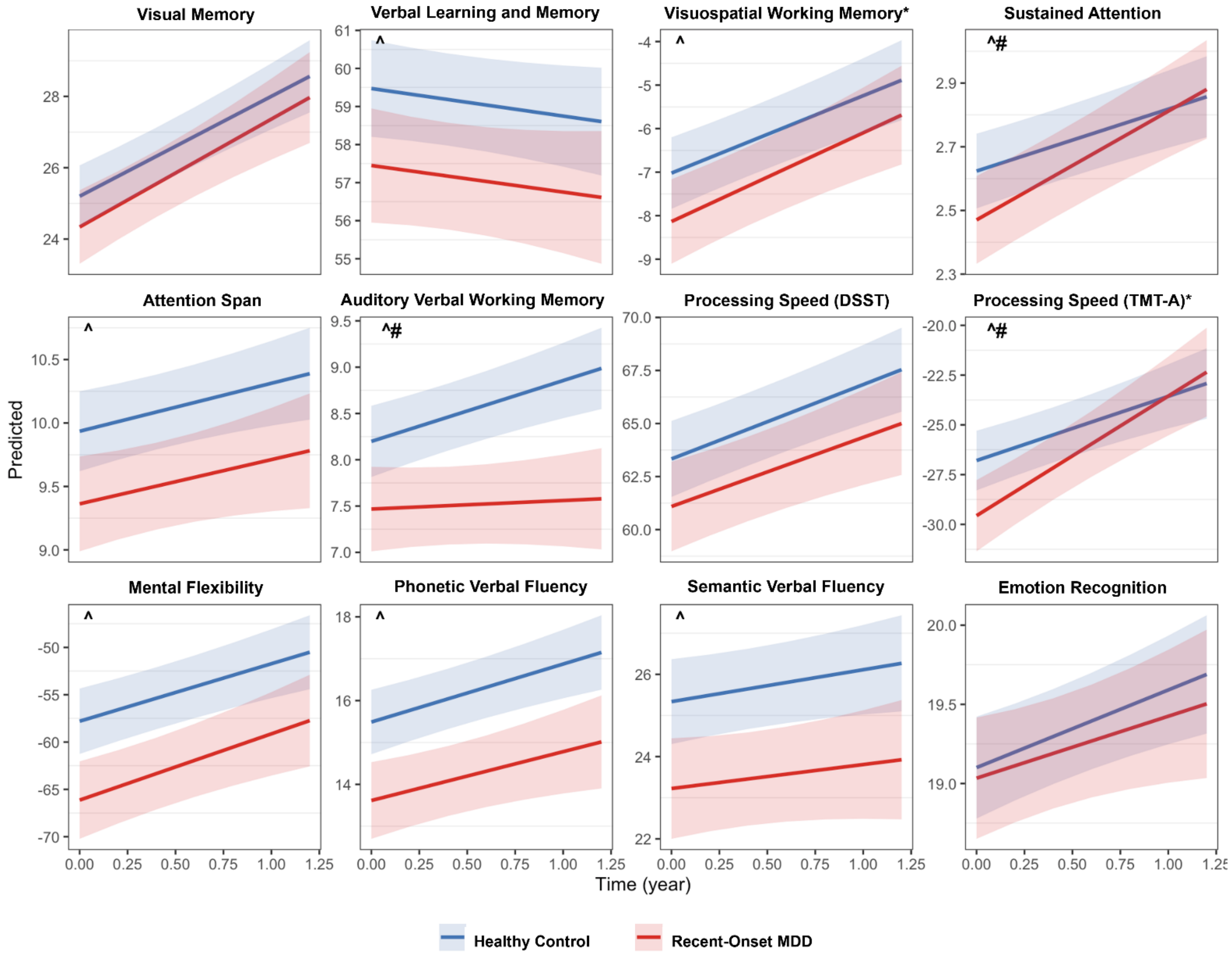


*Reverse-scored so that higher scores indicate better performance. ^Significant variation in intercept. #Significant variation in slope.
